# Antigen-specific tolerization in human autoimmunity: Inhibition of interferon-beta1a anti-drug antibodies in multiple sclerosis

**DOI:** 10.1101/2021.01.24.21250414

**Authors:** Monica Marta, David Baker, Paul Creeke, Gareth Pryce, Sharmilee Gnanapavan, Gavin Giovannoni

## Abstract

**Background:** Antigen-specific tolerance in auto-immune diseases is the goal for effective treatment with minimal side-effects. Whilst this is achievable in animal models, notably via intravenous delivery of the model-specific autoantigen following transient CD4 T cell depletion, specific multiple sclerosis autoantigens remain unproven. However, anti-drug antibodies to human therapeutic proteins represent a model human autoimmune condition, which may be used to examine immune-tolerance induction. Some people with MS (PwMS) on interferon-beta1a (IFNβ1a) develop neutralizing antibodies to IFNβ1a that do not disappear in repeated tests over years.

**Methods:** One PwMS was recruited, as part of a planned phase IIa trial (n=15), who had developed neutralizing antibodies to subcutaneous IFNβ1a. Mitoxantrone (12mg/m^2^) was administered as a lymphocyte depleting agent followed by four days of (88μg/day + three 132μg/day) intravenous IFNβ1a. Subcutaneous IFNβ1a three times a week was maintained during follow-up. IFNβ1a neutralizing antibody responses in serum were measured during treatment and three-monthly for 12 months.

**Findings:** One participant was recruited and, within 6 months of tolerization, the neutralizing antibodies were undetectable. The tolerization treatment was well tolerated. However, the study was terminated after the first enrolment, on ethical grounds, as treatment alternatives became available and the potential risks of mitoxantrone use increased.

**Interpretation:** The data suggest that it may be possible to induce antigen-specific tolerance by providing tolerogenic antigen following transient immune depletion. Further studies are warranted.

**Funding:** The study was supported by an unrestricted research grant from Merck-Serono.

## Introduction

Multiple sclerosis (MS) is an immune-mediated, demyelinating disease of the central nervous system. Although inhibition of relapsing inflammatory disease is possible, such immunotherapy can be associated with side-effects following the non-selective targeting of large parts of the immune system (Marta & Giovannoni 2012). Antigen-specific therapy would offer the benefit to selectively target pathogenic cells and is already and consistently achievable in experimental (auto)immunity via delivery of autoantigens via tolerogenic, notably intravenous, routes following transient T-cell depletion (Polak & Turk 1969; Lanoue et al. 1997, Pryce et al. 2005; Smith et al. 2005). The challenge is to translate this simple combinational approach into human autoimmunity in general and into MS in particular. Although T-cell depleting antibodies succeeded in experimental animals (Polak & Turk 1969; Lanoue et al. 1997; Pryce et al. 2005), the failure of CD4-antibody-mediated depletion in MS (van Oosten et al. 1997) meant that alternative depleting drugs would be required for clinical studies. Mitoxantrone is an immunosuppressive agent that had a role for the treatment of aggressive MS and could be used as a depleting drug to promote tolerance induction (Pryce et al. 2005; Cocco & Marrosu 2014; Martinelli Boneschi et al. 2013). However, the autoantigens in MS are unknown and attempts to induce antigen-specific tolerance via monotherapy have, unsurprisingly, consistently failed (Weiner et al. 1993; Kappos et al. 2000; Garren et al. 2008; Freedman et al. 2011; Lutterotti et al. 2013; van Noort et al. 2015; Lutterotti et al. 2019). Nevertheless, there are autoimmune conditions where the autoantigen is known, and one example is the anti-drug antibody response to a therapeutic human protein. Some people treated with human interferon beta1a (IFNβ1a) develop neutralizing antibody responses that can stop IFNβ1a working (Vallbracht et al 1981; Abdul-Ahad et al. 1997; Farrell et al. 2011). In other autoimmune disorders such as rheumatoid arthritis, Sjögren’s and Crohn’s disease, concomitant methotrexate use attenuates neutralizing antibody frequency, but not all disease-modifying agents have been demonstrated to impact on immunogenicity (Jani et al. 2014). Although there are currently many treatment options for relapsing MS, when this study started, only IFNβ1 and glatiramer acetate were available in the United Kingdom (Marta & Giovannoni 2012). The development of neutralizing antibodies could mean treatment failure (Farrell et al. 2011). This study was designed to test the hypothesis that a combination of lymphocyte depletion with mitoxantrone and intravenous IFNβ1a could reverse IFNβ1a-specific neutralizing antibody response.

## METHODS

### Study Design

The single-centre, open-label phase IIa study was conducted in accordance with the Declaration of Helsinki/International Conference on Harmonisation-Good clinical practice was approved by the Cambridgeshire 4 Research Ethics Committee (REC 08/H0305/64) in 2009. There was no adequate data for sample size calculation. The study planned to recruit n=15 people.

### Registration

EudraCT number 2008-000256-26

### Inclusion criteria

People with clinically-definite MS (according to McDonald 2005 criteria) MS (PwMS), aged 18-65 years and with Expanded Disability Status Scale ≤ 6.5, who had received IFNβ1a (subcutaneous Rebif®-22/44 three times a week or weekly intramuscular Avonex®) for at least 12 months. If they had at least one significant relapse in the prior 12 months and were considering switching therapy, consent was sought. Participants with two consecutive positive neutralizing antibody tests (titre >20U), at least 4 weeks apart, could participate. Male or female participants had to agree to adequate contraception during the study and females had to have a negative pregnancy test at screening.

### Exclusion criteria

People treated with immunosuppressive, immunomodulatory or experimental treatments within the 6 months prior to enrolment, except for steroid use for treatment of relapse or IFNβ1a, were excluded. In addition, participants should not exhibit a history of liver toxicity or abnormal liver function tests (>2.5 times upper limit of normal); poorly controlled diabetes; hypertension; severe cardiac insufficiency, unstable ischaemic heart disease, left ventricular ejection fraction <50% on echocardiogram or an abnormal 12-lead electrocardiogram; complete white blood cell counts <1.5 the lower limit of normal; or have any medical condition which, in the opinion of the chief investigator, would pose additional risk. Participants could not have chronic or recurrent infections or human immunodeficiency virus, a history of malignancy or drug or alcohol abuse and should not be pregnant or breastfeeding.

### Treatment Intervention

Single 12mg/m^2^ intravenous mitoxantrone infusion (Day 0). On day 14, two 44μg intravenous IFNβ1a (Rebif® new formulation) injections (88μg/day), if the first 44µg dose was tolerated. This was followed by four daily intravenous injections of 132μg/day (total dose 660μg). If safe in the initial five participants, the dose would be escalated to a total dose of 1320μg in a second cohort (264μg/day. n=5) and a third cohort (n=5) of 2640μg over 5 days. The maximum dose administered was calculated to limit excessive exposure of bioactive intravenous IFNβ1a after saturation (quenching) of neutralizing antibodies. Participants would stay on their IFNβ1a treatment for the rest of the year-long study.

### Outcome measures

The primary outcome was to assess the therapeutic safety and efficacy of intravenous IFNβ1a after transient peripheral leucocyte depletion. Planned secondary outcomes were to measure T-cell proliferation to IFNβ1a, and T and B cell cytokine responses relative to baseline, compared to response to tetanus toxoid control antigen. Tertiary outcomes were to measure neutralizing antibody titre at 3, 6, 9 and 12 months relative to baseline. Serum neutralizing antibody responses were monitored using a validated luciferase-based reporter neutralizing assay (Lam et al. 2008; Farrell et al. 2011).

### Safety

Adverse events were monitored throughout the trial. Laboratory (Haematology and biochemistry) tests and physical examinations were performed at screening, during drug treatment days and 3-monthly trial visits.

## RESULTS

The trial was planned in 2006 and funding obtained in 2007. Regulatory approval was sought in early 2008 and was granted in early 2009. However, recruitment was delayed whilst accredited neutralizing antibody assays were brought in-house. Recruitment for the trial was slow and only one participant was enrolled by the end of 2012 (Figure 1). The trial was terminated in 2013 due to ethical concerns related to the higher than predicted risk posed by the use of mitoxantrone in MS (Ellis et al. 2013) and other licenced and effective alternatives becoming available. The participant received only four (88μg/day and three 132μg/day) cycles of intravenous IFNβ1a, as they developed grade 4 neutropenia (which subsequently resolved) so the last daily dose was not injected, as per protocol. There were no other relevant serious adverse events. There were 8 adverse events recorded (Supplementary data. Figure 1S). With only one participant, we did not evaluate secondary (immune parameters) outcomes. In the trial participant, the neutralizing antibody levels dropped below the level of detection within 3 months and in following serum samples (Figure 1). Serial analyses of serum samples from 13 people with MS treated with IFNβ1a three times a week over a period of 1 year failed to show any case of reduced neutralizing titres (Figure 1). This supports the view that antigen-specific immune tolerance to IFNβ1a may have been induced.

**Figure 1.**
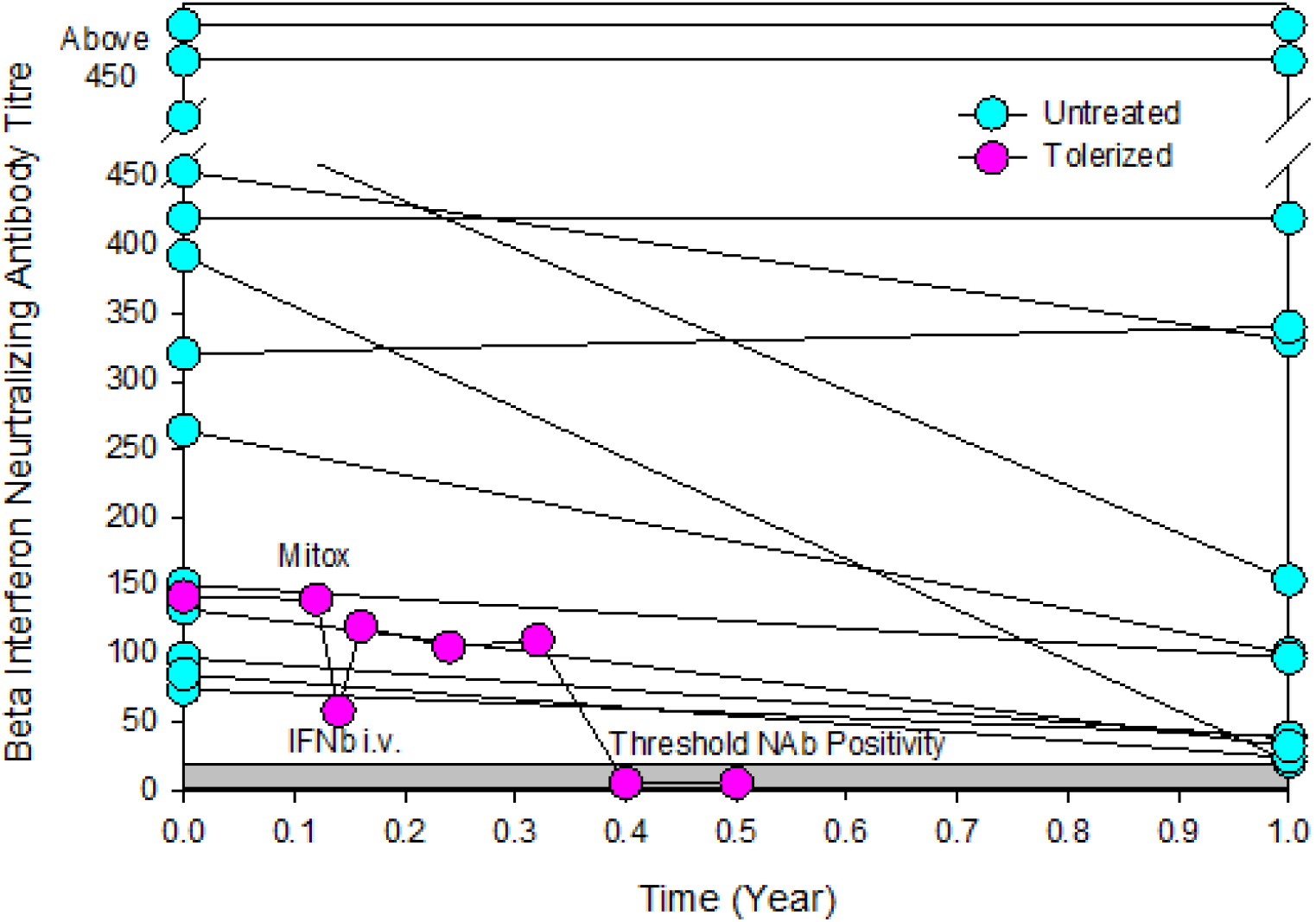
Intravenous *IFNβ1a*-specific tolerance induction inhibited neutralizing antibody responses.

Individual IFNβ1a neutralizing antibody (Nab) responses at baseline and 1 year later in people treated with 44µg IFNβ1a subcutaneously three times a week (n=13) and the response in the trial participant receiving an infusion of mitoxantrone (Mitox) followed four days of intravenous IFNβ1a.

## DISCUSSION

This case-study suggests that it may be possible to induce antigen-specific tolerance to proteins in humans. The combination of lymphocyte depletion with mitoxantrone followed by intravenous high-dose antigen, eliminated autoimmune antibody responses and thus contrasts with the lack of efficacy following high-dose intravenous IFNβ1 monotherapy (Hegen et al. 2014). Whilst our study lacked supportive mechanistic in vitro data, due to early trial termination, varied mechanisms of intravenous tolerance have been well studied in animals and humans (Smith et al. 2005; Lutterotti et al. 2013). Although intravenous tolerance develops within hours (Pryce et al. 2005), immunoglobulin half-life is about 3-4 weeks (Waldmann & Strober 1969), which is consistent with the decrease in titre of neutralizing antibodies over months. Although promising, the trial was terminated early. This was because an oral, higher efficacy, alternative drug became available that could be used following treatment failure with IFNβ1 (Polman et al. 2006; Weissert 2006). Notably fingolimod was approved for use in the United Kingdom in 2013 and was more attractive than continuing on frequent lower-efficacy, injectable therapies. Natalizumab was approved for use in the United Kingdom National Health Service but not widely available as the use was restricted to rapidly evolving, severe, relapsing MS. In addition, prior mitoxantrone use increased the risk for progressive multifocal leukoencephalopathy during natalizumab treatment (Faulker 2015). During this period, mitoxantrone was also reported to cause worse fertility complications (Cocco et al. 2008) and importantly, studies emerged showing that the risk of mitoxantrone-induced leukaemia had doubled (Ellis & Boggild 2009; Ellis et al. 2015). Therefore, even if we hypothesized that treatment could reduce infection risks associated with inhibition of endogenous IFN intrinsic pathways (Fine et al 2014), it was not ethically justifiable to expose people to the procedure and thus the planned trial became an anecdotal case report.

It is possible that loss of the neutralization response was unrelated to treatment, but this would be unlikely, as it was not found in non-tolerized individuals and those given intravenous IFNβ1a alone (Hegen et al. 2014). Monotherapy, intravenous tolerance continues to be trialled and repeatedly found to be insufficiently successful in MS (Freedman et al. 2011; Hegen et al. 2014; Lutterotti et al. 2013; van Noort et al. 2015), as would be predicted from extensive animal studies, even if the correct autoantigens were selected (Lanoue et al. 1997; Pryce et al. 2005). These studies indicate the essential need to deplete pathogenic cell subsets prior to tolerance induction in established disease and thus casts doubt on the value of tolerogenic monotherapy without prophylactic lymphocyte depletion (Lanoue et al. 1997; Pryce et al. 2005; von Kutzleben et al. 2017; Krienke et al. 2021). Although neutralizing responses to other therapeutic proteins can be a problem in MS (Dubuisson et al. 2018; Baker et al. 2020), given the variety of current treatments (Dobson and Giovannoni 2019), MS is perhaps not the best disease to develop this approach further. However, targeting neutralizing responses to antibodies that provide life-saving function (Patriquin & Kuo 2019) or autoimmune disease with a defined autoantigen and limited treatment options may be a better target for future studies.

## Data Availability

DATA SHARING STATEMENT: Anonymous data is available on request

## ACKNOWLEDGEMENTS

The authors would like to thank Forian Deisenhammer and Mike Boggild and for providing advice on the intravenous delivery of IFNβ1a and mitoxantrone risk assessment.

## AUTHOR CONTRIBUTION

Concept: DB, GP, MM, GG, Initial Draft: DB, MM; Assays: PC; Study recruitment: MM, SG, GG; Study funding: GG; Regulatory Issues: GG, SG, MM; Trial conduct MM, GG; Final manuscript: All

## DATA SHARING STATEMENT

Anonymous data is available on request

## Supplementary Data

**Figure 1S.**
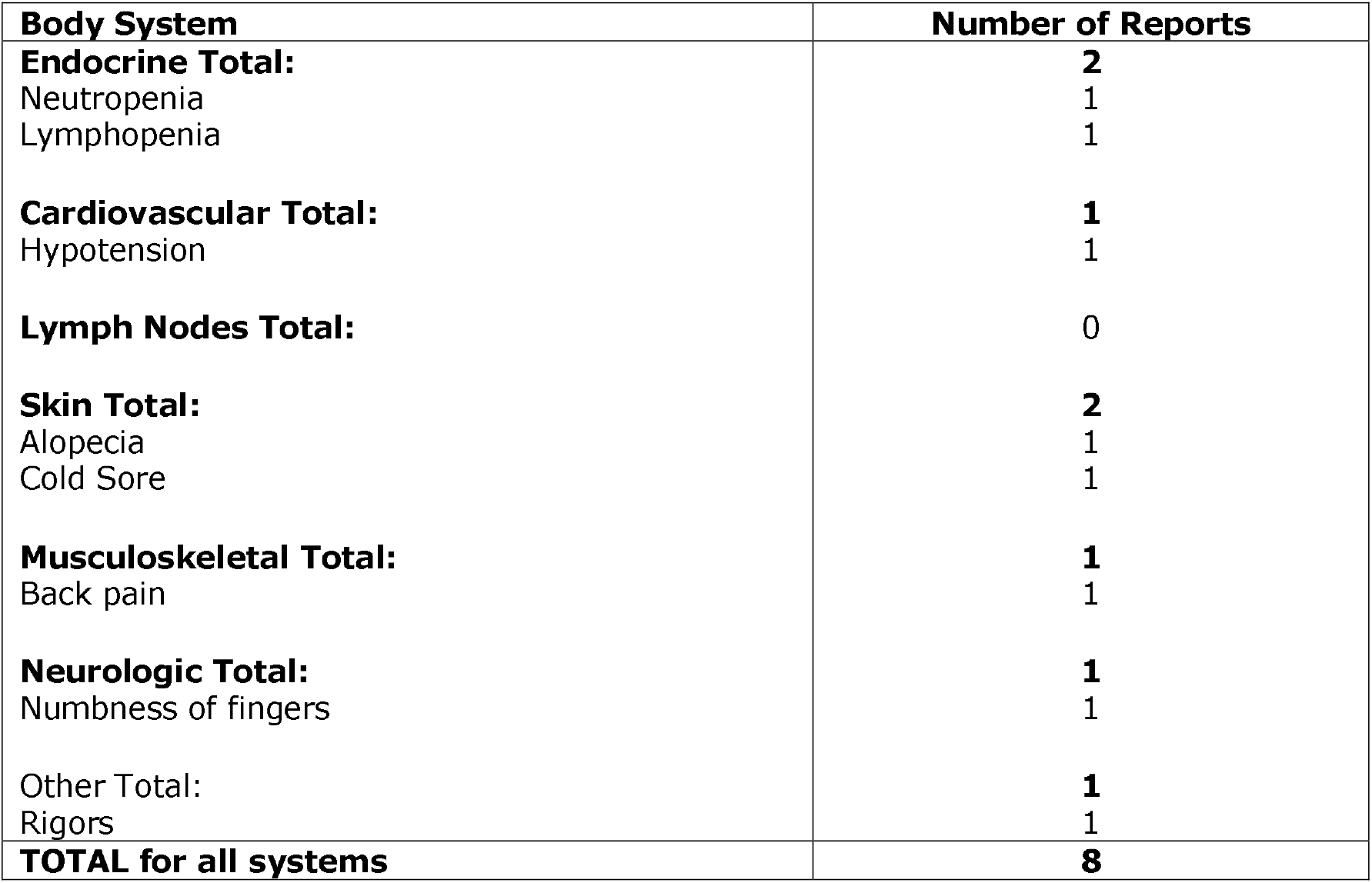
Adverse events by Body system for the duration of the trial

